# Impact of Kaposi Sarcoma on Quality of Life Amongst HIV-infected Adults Initiating Antiretroviral Therapy in East Africa

**DOI:** 10.1101/2023.07.21.23292658

**Authors:** Scott Lu, Miriam Laker-Oketta, Helen Byakwaga, David Glidden, Bwana Mwebesa, Conrad Muzoora, Toby Maurer, Melissa Assenzio, Peter Hunt, David Bangsberg, Jessica Haberer, Jeffrey Martin

## Abstract

**BACKGROUND:** In sub-Saharan Africa, increased antiretroviral therapy (ART) availability has improved survival after diagnosis of Kaposi sarcoma (KS) compared to the pre-ART era, but mortality among patients with KS is still considerably higher than HIV-infected persons without KS. Furthermore, among those patients with KS who are treated initially with ART without adjunct chemotherapy and who do survive, little is known about how well they function and feel — quality of life (QOL) — compared to those without KS.

**METHODS:** Among HIV-infected adults initiating ART in two prospective studies in Uganda, we compared those presenting with KS to those without KS. QOL was measured using the Medical Outcomes Survey-HIV instrument prior to ART initiation and at 16, 32, and 48 weeks thereafter; higher scores indicate better QOL. To ascertain the independent effect of KS versus non-KS on 11 domains of QOL and two summary scores, we created mixed effects models adjusted for directed acyclic graph-informed confounders.

**RESULTS:** We examined 224 participants with KS and 730 without KS, among whom 64% were women and median age was 34 years. Prior to ART initiation, participants had a median CD4+ T count of 159 cells/mm^3^ and plasma HIV RNA of 5.1 log_10_ copies/ml. In adjusted analyses prior to ART initiation, those with KS had lower mean scores in 8 of 11 QOL domains and both physical and mental health summary scores compared to those without KS. After 48 weeks of ART, those with KS had higher mean QOL scores compared those without KS in 4 domains and the mental health summary score, and lower scores in only one domain. There was no significant difference in 6 domains and the physical health summary score,

**CONCLUSIONS:** Amongst HIV-infected adults in East Africa, at time of ART initiation, those with KS had worse mean QOL compared to those without KS. Over the first year of ART, those with KS became comparable to or exceeded those without KS in most QOL domains. The findings indicate that some patients with KS can be treated with ART alone and further emphasize the need to predict those who will do well with ART alone versus those who need additional initial therapy.

## Introduction

In sub-Saharan Africa, despite widely increased access to antiretroviral therapy (ART), HIV-associated Kaposi’s sarcoma (KS) remains among the most commonly occurring cancers in the region (1), and the most common overall cancer in some countries (2, 3). In 2020, GLOBOCAN estimated that KS was the 5th most common cancer among men in sub-Saharan Africa and 11th most common in women (4) (and these counts are likely substantial underestimates (5). While not eliminating the occurrence of KS, increased accessibility to ART in Africa has, where data are available, improved survival after diagnosis of KS. In the pre-ART era, mortality at one year following KS diagnosis ranged from 55 to 70% (6). Following the rollout of ART, one-year mortality is now varying from 22 to 28% (7-9).

Although survival amongst those with HIV-related KS in sub-Saharan Africa has improved in the ART era, mortality remains considerably worse compared to HIV-infected patients without KS. For example, one report from South Africa found that patients with KS had four times the rate of death compared to those without KS in the first year of ART (10). It is hypothesized that some of this decrement in survival is explained by the use of ART alone as initial therapy (i.e., without concurrent chemotherapy), a practice motivated by successes of ART alone in small series of patients in resource-rich settings (11) but never formally evaluated in sub-Saharan Africa. Furthermore, among those patients with KS who are treated initially with ART alone and are able to survive, little is known about how they function and feel — their quality of life (QOL) — compared to those without KS. This is relevant because KS can cause significant morbidity, both physically from the tumor burden and psychosocially from the distinctive skin lesions. To the extent that ART alone can normalize QOL is a critically important aspect of the effectiveness of ART alone as initial therapy for KS.

To address the effect of initial treatment of KS with ART alone on QOL, we identified adults living with HIV initiating first-time ART in Uganda and compared those with KS to those without KS. We evaluated QOL, measured in several domains, prior to starting ART and in the year following ART initiation. This allowed us to ask how having KS, compared to not, impacts QOL given survival in the first year after starting ART. The findings add to the evidence base regarding the appropriateness of ART alone as the initial therapy for KS in sub-Saharan Africa.

## Methods

### Overall design

HIV-infected adults in Uganda with KS and without KS, who were initiating first-time ART, were compared regarding a variety of QOL measures over 48 weeks. The first objective was to define the effect of KS on QOL in the natural history state, prior to ART. The second objective was to determine to what extent does initial treatment of KS with ART alone improve QOL over time and approximate the QOL experienced by HIV-infected patients without KS.

### Participants

We combined data from two linked prospective studies in Uganda. The Antiretrovirals in Kaposi Sarcoma (ARKS) study included participants with KS, and the Uganda AIDS Rural Treatment Outcomes (UARTO) cohort included the participants without KS. ARKS was a randomized trial comparing the effect of a protease inhibitor (PI)-based ART regimen (lopinavir/ritonavir plus emtricitabine/tenofovir) to a non-nucleoside reverse transcriptase inhibitor (NNRTI)-based ART regimen (efavirenz plus emtricitabine/tenofovir). Eligible participants were ART-naïve HIV-infected adults (≥ 18 years) with newly diagnosed KS who were deemed to have no functional impairments that necessitated immediate chemotherapy and thus were initially treated with ART alone. All diagnoses of KS were either biopsy-confirmed or cases limited to lesions highly suggestive of KS but restricted to the oral cavity and not amenable to biopsy. Patients with active, untreated opportunistic infection or other malignancies were excluded. The fieldwork was based in Kampala, Uganda, but recruitment occurred from throughout Uganda. Participants were followed for 48 weeks. The study found no significant difference between the randomization groups, and thus all participants from ARKS were pooled for the current analysis.

UARTO enrolled a consecutive sample of ART-naïve HIV-infected adults who were attending the Immune Suppression Syndrome Clinic at the Mbarara Regional Referral Hospital in Mbarara, Uganda and had indications for starting ART as deemed by local providers and ambient treatment guidelines. The predominant ART regimen used was NNRTI-based. For the current analysis, all participants who were diagnosed with KS either at the baseline visit or at the first follow-up visit at 4 months (and thus may have had undocumented KS at baseline) were excluded. Participants were followed for up to 7 years, but only the first 48 weeks were assessed in the current analysis. UARTO was developed in part to provide a comparator for ARKS and as explained below, utilized identical measurements and the same data coordinating center.

All participants provided written informed consent.

### Measurements

#### Sociodemographic, economic and clinical characteristics

Prior to starting ART, participants were asked, using interviewer-administered questionnaires, about demographic characteristics, literacy, employment status, income, asset holdings via the Filmer-Pritchett index (12), and medical history.

#### Quality of life

The Medical Outcomes Study HIV Health Survey (MOS-HIV) was designed to assess functional status and well-being among HIV-infected persons (13). It consists of 35 questions covering 11 domains (general health perceptions, pain, physical functioning, role functioning, social functioning, mental health, energy, health distress, cognitive functioning, quality of life, and health transition). Each of the 11 domains is summarized in a score ranging from 0-100, with higher values indicating better QOL. Separate overall physical health summary (PHS) scores and overall mental health summary (MHS) scores are also derived, also on a scale for 0 to 100 (14). We used a version of the MOS-HIV translated to the local language (Luganda) in Uganda and culturally adapted (15, 16). All participants completed the survey at the baseline visit just prior to starting ART. Among those with KS in ARKS, the survey was completed every 4 months thereafter. Among those without KS in UARTO, the survey was administered every 3 months in the early portion of the study and every 4 months among later enrollees.

#### Vital status

In both ARKS and UARTO, intensive efforts were made to update participant vital status if participants failed to return for research visits. This included prospective collection of residence location information and extensive tracking in the community.

#### Laboratory

Plasma HIV RNA level was measured using the Amplicor HIV Monitor version 1.5 or the Cobas TaqMan HIV-1 version 1.0 assays (Roche, Branchburg, NJ). CD4+ T cell counts were assessed using the FACSCalibur™ system (Becton Dickinson, San Jose, CA). All testing was performed at the same laboratory at the Infectious Diseases Institute in Kampala.

### Statistical analysis

The independent effect of having KS on QOL both prior to ART initiation and, among those still alive, at several time points during the first 48 weeks of ART was determined with mixed effects linear regression. Adjustment for confounding was informed by a directed acyclic graph (Figure X) (17, 18). Participants with KS were compared to those without KS for mean QOL scores in the aforementioned 11 domains as well as the physical summary score and mental summary score. Participants not known to have died but yet failed to attend scheduled follow-up visits had their QOL measurements imputed via multiple chained imputation (19), based on an assumption of missing at random conditional on age, biological sex, body mass index (BMI), Filmer-Pritchett asset index, literacy, employment status, income, history of cryptococcus tuberculosis, CD4+ T cell count, plasma HIV RNA level, vital status, and all available prior measurements of QOL. The differential mortality can induce selection bias in QOL when QOL and survival share unmeasured common causes. We conducted sensitivity analysis using a principal stratum approach comparing the possible QOL difference in the latent subset of those who would survive with or without KS (20). In both the mixed effects regression and the multiple imputation, continuous variables were specified with restricted cubic splines. Vital status variables included a dichotomous variable for known mortality, a continuous variable indicating duration of time before the end of follow-up or mortality, and an interaction term including both. The difference in mean QOL values (and 95% confidence intervals) between participants with and without KS, adjusted for relevant confounding, was expressed with marginal estimates integrating over the range of covariates for the entire study population using the ‘margins’ command in Stata (version 16; Stata Corporation, College Station, Texas).

## Results

### Characteristics of participants at time of ART initiation

A total of 954 participants were evaluated from the time of ART initiation, including 224 individuals newly diagnosed with KS and 730 individuals not known to have KS. The distribution of age was approximately the same in the KS and non-KS groups (median 34 years and 33 years, respectively), but the KS group had fewer women (44%) than the non-KS group (70%) (Table 1). Both groups featured a sizeable fraction of participants with only primary school-level of education and less than complete literacy. Participants in the KS group had a lower median CD4+ T cell count (111 versus 173 cells/mm^3^), but the distribution of plasma HIV RNA content was approximately the same (median 5.2 compared to 5.0 log_10_ copies/ml). Regarding ART, those in the KS group were all administered efavirenz-based therapy while in the non-KS group, nevirapine- and efavirenz-based therapies were equally used.

### Effect of KS on QOL prior to ART initiation

To understand the influence of KS on QOL in the natural history state, we compared QOL in the KS group versus the non-KS group just prior to ART initiation. Before adjusting for confounders, median PHS and MHS scores among those with KS were lower than those without KS. Regarding individual QOL domains, participants with KS had lower median scores in 7 of 11 domains: health distress, mental functioning, pain, quality of life, role functioning, social functioning, and health transition (Table XX). After adjustment for sex, BMI, education, income, Filmer-Pritchett asset index, plasma HIV RNA level, and CD4+ T cell count, those with KS had significantly lower mean PHS and MHS scores than those without KS (Figure X and T). Regarding the individual QOL domains, participants with KS had significantly lower mean scores in 8 of 11 domains: pain, general quality of life, role functioning, social functioning, energy, mental functioning, health distress, and health transition. There was no strong evidence of a difference between groups in the mean scores in the domains of general health perception, cognitive function, and physical function.

### Effect of KS on QOL following ART initiation

After initiation of ART, vital status was ascertained for all participants in the KS group and 99% of the non-KS group at 48 weeks of follow-up. Throughout follow-up, 56 (25%) of the KS group had developed an indication for chemotherapy, of which 52 were prescribed a chemotherapeutic agent. By 48 weeks, 44 (20%) participants in the KS group and 41 (6.0%) participants in the non-KS groups had died. Among those participants not known to have died, 777 of 784 (99%) expected study visits were completed (with assessment of QOL) in the KS group and 2,764 of 3130 (88%) in the non-KS group. QOL data were imputed when visits were not completed.

In the summary scores and virtually all eleven individual domains, both those with and without KS experienced gains in QOL scores after ART initiation (Figure X and Y and Table Z). In general, there were improvements in QOL scores early after ART initiation with a subsequent plateau around 16 weeks in the non-KS group while the KS group had monotonic increases. After adjustment for the same factors detailed in the pre-ART analysis above, there was no strong evidence at 48 weeks for a difference between the KS and non-KS groups in the mean PHS score but evidence for higher mean score in the KS group for the MHS score (Figure X and T). Regarding the individual QOL domains, there was no strong evidence at 48 weeks for a difference between the groups in the means scores in 4 of 11 domains (pain, social functioning, health distress, and physical functioning) and evidence for higher mean scores among the KS group in 6 of 11 domains (general health perceptions, general quality of life, energy, mental functioning, cognitive functioning, and health transition). It was only in the domain of role functioning in which, despite narrowing the gap since baseline, those with KS persisted with lower mean scores. No significant differences were found via principal stratum analysis.

## Discussion

As much as any cancer, KS has substantial potential to affect QOL. Patients with KS almost always have cutaneous lesions (ref), which have the potential to be in view of the affected individual and others; cosmetic manifestations can be substantial (21). Furthermore, while KS lesions are often painless, they can ulcerate, become superinfected, and be associated with lymphatic obstruction and edema. As such, any study of the impact or treatment of KS merits evaluation of QOL. Among HIV-infected adults in East Africa who were about to begin ART, we not surprisingly documented that those with KS had poorer mean QOL scores on virtually all aspects of the well-validated MOS-HIV survey. After beginning ART, we found that, among patients who survived, both those with and without KS had improvements in their scores of most QOL domains over the first 48 weeks of therapy. Notably, those with KS reported mean QOL scores that either exceeded or equaled those without KS in almost all domains. As the majority of the KS group did not receive chemotherapy, it can be argued use of ART alone as initial therapy for KS abrogated the gap in QOL. Although this work did not feature a direct comparator to ART, the findings lend insight as to the appropriateness of ART alone as the initial therapy for HIV-related KS in sub-Saharan Africa. Specifically, at least some patients with HIV-infected KS, but not all, can be successfully treated with ART alone.

In the natural history state (prior to an individual’s use of ART), we found that patients with HIV-related KS had lower PHS and MHS scores compared to non-KS patients. In another study of the effect of KS on QOL, Harris et al. compared HIV-infected men with KS to HIV-infected men without KS using two different types of quality-of-life measurement (22). One method included a time trade off technique (TTO), presenting participants with two sets of hypothetical clinical scenarios, depicting two different qualities of life, to determine willingness to exchange survival time for health state with less severe or ideal health. Their other method asked participants how they would rate each health state on a visual analogue scale from 0 (death) to 100 (best possible health). From a TTO standpoint, they found those with KS were willing to give up more life expectancy of stable disease for a state of the best imaginable health compared to those without KS. They did not note a difference between groups from the health rating system.

Surprisingly few studies have evaluated QOL among patients with HIV-related KS in resource-limited settings who undergo therapy for KS. The Kaposi sarcoma AIDS AntiRetroviral Therapy (KAART) study in South Africa randomized treatment-naïve HIV-infected adults with KS to receive ART or ART combined with chemotherapy (23). KAART demonstrated improvements in global health status, emotional and cognitive functioning, fatigue, pain, insomnia, constipation, diarrhea, and financial problems among both treatment arms; improvement in pain and overall quality of life was greater in those receiving chemotherapy compared to the ART treatment arm. KAART noted worsening role functioning, though the specific items assessing this domain differed between their scale (24) and the MOS-HIV. Our results contrast starkly with the findings of Olweny et al. who compared chemotherapy and radiotherapy to supportive care in Zimbabwe between 1994 and 1999, before the advent potent ART (6). Olweny et al. concluded etoposide, Actinomycin-D (ABV, radiotherapy, and supportive care all resulted in decreasing QOL using the functional living index-cancer (FLI-C) and a novel KS-specific instrument they developed. These results are likely explained by the absence of ART and the inferior potency of the ambient interventions (25). Of note, we are unaware of any work similar to ours that compared QOL after treatment to a counterfactual state of not having KS, in other words, evaluating just how close to “normal” QOL can be achieved with therapy.

Some limitations in our work should be considered. Regarding selection bias, missing QOL data following start of ART is the principal threat but one we have mitigated with multiple imputation (26). Furthermore, the number of missing QOL datapoints were relatively few. In the realm of measurement, we consider the most robust approach to QOL analysis includes a general QOL instrument combined with a disease-specific module (27). Because there is no disease-specific QOL instrument available for KS, we were limited to a general tool (MOS-HIV) and thus may have lacked sensitivity to detect differences between those with KS and those without. Relatedly, unadjusted and adjusted analyses both demonstrated an apparent increase in mean QOL scores in several mental health and social functioning domains among those with KS relative to those without. This seemingly implausible finding might tend to discredit the QOL measurements, but alternatively we hypothesize that because QOL scores in these domains started at much lower mean levels among those with KS, their improvements may have been perceived as more remarkable (and hence reported more favorably) than comparable improvements in those without KS. Lastly, regarding confounding, the observational nature of the analysis leaves open the possibility of unmeasured confounding when comparing those with KS to those without. We believe our robust measurement of and adjustment for a number of socioeconomic, clinical, and laboratory parameters largely mitigates unmeasured confounding but cannot definitively dismiss it.

Although not a threat to the internal validity of our results, our findings may not be generalizable to those with advanced KS who require chemotherapy in addition to ART as initial therapy. In particular, many patients with visceral disease, oral lesions which interfere with chewing or swallowing, or symptomatic KS-related edema may plausibly not normalize their QOL; their disease may be too severe for currently available treatment to impact. In particular, KS-related edema is anecdotally known not to readily resolve with therapy (28) and barring patient’s accommodation to the manifestations, it would not be expected for QOL to normalize. These speculations emphasize the importance of including QOL assessment in all treatment studies of HIV-related KS. That is, improvement in survival is not tantamount to improvement in QOL in all surviving patients.

A combination of factors have led to many patients with HIV-related KS in sub-Sahara Africa being treated initially with ART alone. Lack of affordable chemotherapy is undoubtedly a major reason, but historical influences are also important. By the time ART became widely available in sub-Saharan Africa, ART alone in resource-rich settings had been anecdotally reported to be effective in the treatment of KS in small case series (29-33). These reports and lack of international guidelines essentially gave sub-Saharan clinicians permission, if not encouragement, to use ART alone for patients with HIV-related KS in resource-limited settings when ART finally became available. The aforementioned KAART study (23) fueled this enthusiasm when finding no strong evidence for a difference in overall survival or QOL between those treated with ART alone vs ART plus chemotherapy. (Of note, the large fraction of patients randomized to chemotherapy but who ultimately did not receive chemotherapy may have resulted in an overly optimistic view of the effectiveness of ART alone.) Subsequently, formal treatment recommendations from both the World Health Organization (34) and National Comprehensive Cancer Network (35) allowed for ART alone in the case of asymptomatic mild/moderate local cutaneous (T0 or Stage I/II) disease. Collectively, the available data suggests that ART alone is better than no therapy, equivalent to ART plus modestly potent chemotherapy in some patients, and inferior, at least in some groups, to ART plus the most effective chemotherapy (liposomal doxorubicin or paclitaxel). Stated another way, there are some patients with HIV-related KS who can be treated with ART alone and have equivalent survival to what would occur with addition of chemotherapy. The value of our current work is that it indicates that not only do some patients with HIV-related KS who are treated with ART alone survive but they also normalize QOL in reference to the most relevant contrast, persons with HIV disease who are without KS. However, knowing that some patients can have favorable outcomes with ART alone when other more effective (but more expensive) interventions are available is not useful clinically. Accordingly, we believe the most salient question in the area of treatment for KS in sub-Saharan Africa is predicting which patients can be successfully treated with ART alone. By prediction, we mean more than just discovering a set of risk factors for favorable outcomes. Rather, we mean development of a clinical prediction rule that can precisely estimate probability of success with initial treatment with ART alone, thus giving patients and providers more objective information as to whether to seek more expensive therapies.

In conclusion, among HIV-infected patients in East Africa, we documented the influence of KS on QOL. The diminution of QOL among those with KS in the untreated state is broad in spectrum. Treatment with ART alone — in this study population that had no immediately life-threatening complications of KS — closed the gap in QOL between those with KS and those without in virtually all domains of QOL. The findings indicate that some patients with KS can be successfully treated with ART alone and further emphasize the need to predict those who will do well with ART alone versus those who need additional initial therapy.

## Data Availability

All data produced in the present work are contained in the manuscript.

## Notes

**Funding:** Supported by National Institutes of Health (U54 CA190153 and U54 CA254571)

### Competing Interest Statement

The authors have declared no competing interest.

### Funding Statement

Supported by National Institutes of Health (U54 CA190153 and U54 CA254571)

### Author Declarations

IRB of UCSF gave ethical approval of this work

